# Diagnosis of SARS-CoV-2 infection with LamPORE, a high-throughput platform combining loop-mediated isothermal amplification and nanopore sequencing

**DOI:** 10.1101/2020.09.18.20195370

**Authors:** Leon Peto, Gillian Rodger, Daniel P Carter, Karen L Osman, Mehmet Yavuz, Katie Johnson, Mohammad Raza, Matthew D Parker, Matthew D Wyles, Monique Andersson, Anita Justice, Alison Vaughan, Sarah Hoosdally, Nicole Stoesser, Philippa C Matthews, David W Eyre, Timothy EA Peto, Miles W Carroll, Thushan I de Silva, Derrick W Crook, Cariad M Evans, Steven T Pullan

## Abstract

LamPORE is a novel diagnostic platform for the detection of SARS-CoV-2 RNA that combines loop-mediated isothermal amplification with nanopore sequencing, which could potentially be used to analyse thousands of samples per day on a single instrument. We evaluated the performance of LamPORE against RT-PCR using RNA extracted from spiked respiratory samples and from stored nose and throat swabs collected at two UK hospitals. The limit of detection of LamPORE was 7-10 genome copies/µl of extracted RNA. This is above the limit achievable by RT-PCR but was not associated with a significant reduction of sensitivity in clinical samples. Positive clinical specimens came mostly from patients with acute symptomatic infection, and among these LamPORE had a diagnostic sensitivity of 99.1% (226/228 [95% CI 96.9–99.9%]). Among negative clinical specimens, including 153 with other respiratory pathogens detected, LamPORE had a diagnostic specificity of 99.6% (278/279 [98.0–100.0%]). Overall, 1.4% (7/514 [0.5–2.9]) of samples produced an indeterminate result on first testing, and repeat LamPORE testing on the same RNA extract had a reproducibility of 96.8% (478/494 [94.8–98.1]). This indicates that LamPORE has a similar performance to RT-PCR for the diagnosis of SARS-CoV-2 infection in symptomatic patients, and offers a promising approach to high-throughput testing.

## Background

Rapid, reliable and high-throughput methods of testing for SARS-CoV-2 infection would help to control transmission. Present diagnosis relies mostly on RT-PCR, but this has proven difficult to expand to the scale needed for population-wide testing of symptomatic individuals. For example, shortages of laboratory RT-PCR capacity still limit the United Kingdom testing program more than seven months after the SARS-CoV-2 pandemic was declared a public health emergency of international concern by the WHO. Further expansion of testing to include screening of asymptomatic individuals, which may be needed to prevent SARS-CoV-2 circulation, would require a significant further increase in testing capacity^1,2^.

In the UK, clinical laboratories have struggled to expand conventional RT-PCR workflows to meet the demand for SARS-CoV-2 testing, and many have explored alternative methods that would be more scalable or allow near-patient use^3,4^. At the Oxford University Hospitals NHS Foundation Trust (OUH) and Sheffield Teaching Hospitals NHS Foundation Trust (STH), we evaluated LamPORE, a novel diagnostic platform for SARS-CoV-2 that combines loop-mediated isothermal amplification (LAMP) with nanopore sequencing^5^. During sample preparation a unique combination of DNA barcodes is incorporated into the LAMP products from each specimen so that these can be pooled into a single sequencing run. The protocol currently allows up to 96 samples to be analysed on one flow cell, potentially allowing thousands of samples to be analysed per day on a single instrument. The workflow involves a 40-minute amplification, followed by a library preparation and a 60-minute sequencing run, generating results in a comparable time to RT-PCR when starting with extracted RNA.

As well as molecular barcoding, using sequencing to detect the outcome of the LAMP reaction offers other advantages compared with simpler LAMP assays that detect the presence of DNA synthesis by measurement of pH, turbidity, or fluorescent dyes. Sequenced reads from a specific target will contain sequences not present in the primers, avoiding false positives caused by non-specific amplification^6^. Conversely, reads confidently assigned to SARS-CoV-2 target can indicate a true positive even if present at relatively low levels, potentially improving the low sensitivity seen in several LAMP assays compared to RT-PCR^7^. LamPORE uses standard Oxford Nanopore Technologies (ONT) flow cells compatible with several sequencing instruments, including the portable MinION device, and high-throughput GridION and PromethION platforms, so could potentially be used both for mobile and centralised testing.

In this evaluation we aim to compare the performance of LamPORE, which is awaiting regulatory approval, with RT-PCR on extracted RNA from respiratory specimens. Initially, we use spiked samples to determine the analytical limit of detection of the assay. We then use stored clinical samples to determine the assay’s diagnostic sensitivity, specificity and reproducibility.

## Methods

The evaluation was conducted across three sites: OUH, STH and the Public Health England National Infection Service at Porton Down (PHE Porton Down).

### LamPORE

LamPORE is described in detail in James, et al.^5^, and was performed identically at each site using a GridION instrument with operators blinded to sample identity. The assay takes 20µl RNA input into a single multiplex reaction targeting three regions of the SARS-CoV-2 genome; ORF1a, envelope and nucleocapsid genes, plus human β-actin mRNA as a control of sampling adequacy and assay performance. LamPORE sample preparation uses a 96-well plate format, with each sample having one of eight LAMP Forward Inner Primer (FIP) barcodes and one of 12 transposase (rapid) barcodes added before pooling. In these experiments a single LAMP barcode (FIP7) was not used, as it had previously been associated with lower β-actin read counts and was awaiting replacement (unpublished data). As a result, plates contained up to 80 samples, plus two no-template controls and two positive controls consisting of synthetic SARS-CoV-2 RNA (Twist Bioscience).

We used the LamPORE protocol dated 1^st^ July 2020 (version 1, revision 4), the full text of which is available in the supplementary material. Briefly, this consists of adding sample RNA to LAMP master mix and primers, then incubating at 65-80°C in a thermocycler for 40 minutes, during which time amplification occurs and the LAMP primer barcodes are incorporated into concatemers containing the target sequence. Following this, a second set of barcodes are incorporated using a rapid transposase-based method and samples are pooled into a single sequencing library. The pooled library has a bead-based cleanup, then is added to a MinION flow cell and sequenced for 60 minutes, after which a report is generated automatically within seconds for each barcode set. Unlike RT-qPCR, LamPORE is not designed to be a quantitative assay, as measurement only occurs after amplification is complete. The number of reads assigned to each target is used to generate a report as follows:

### Invalid

<50 classified reads in total detected from SARS-CoV-2 and β-actin targets

### Positive

≥50 SARS-CoV-2 reads detected (adding read counts across all three SARS-CoV-2 targets)

### Inconclusive

not invalid and ≥20 and <50 SARS-CoV-2 reads detected

### Negative

not invalid and <20 SARS-CoV-2 reads detected

#### Spiked samples – PHE Porton Down

Spiked samples were prepared and analysed at PHE Porton Down to establish the limits of detection of LamPORE. Aliquots of pooled volunteer saliva were used for spiking experiments, which were confirmed SARS-CoV-2 negative by RT-PCR. These were spiked with cultured SARS-CoV-2 (Victoria/01/202026 passaged twice in Vero/hSLAM cells) and serially diluted with the remaining material to create a dilution series of positive samples.

From each spiked sample a 140µl aliquot was inactivated by addition to 560µl buffer AVL (Qiagen), incubated at ambient temperature for 10 minutes, then added to 560µl 100% molecular grade ethanol. The entire inactivated volume was then extracted manually using the QiaAMP Viral RNA mini kit (Qiagen), with RNA elution into 50µl of nuclease-free water.

For the quantitation of SARS-CoV-2 by RT-PCR, 5µl of RNA extract was used in each of duplicate reactions using the CDC NS1 assay^8^ and the TaqPath 1-Step RT-qPCR Master Mix (Thermo Fisher). All samples were run in a 96-well plate format on a QuantStudioTM 7 Flex System and quantified by comparison to a standard curve of a plasmid 2019-nCoV_N positive control (Integrated DNA Technologies).

#### Clinical specimens – OUH and STH

Testing of stored clinical samples was performed at OUH and STH. All samples were nose and/or throat swabs collected into viral transport media during routine clinical care and stored at -80°C.

#### Sample selection

##### SARS-CoV-2 positive samples

At OUH sequentially available positive samples were chosen without reference to RT-PCR cycle threshold (Ct) value. These were collected from March-April 2020, during which time the PHE RdRp RT-PCR assay was in use^9^, and testing was mostly restricted to hospitalised patients and symptomatic staff. At STH a stratified random sample of specimens collected from April-May 2020 were selected based on their initial SARS-CoV-2 E gene Ct value, with 50% <30 and 50% ≥30. During this collection period, testing at STH was also largely restricted to hospitalised patients and symptomatic staff, using an in-house assay based on the Corman *et al*. protocol^10,11^.

##### SARS-CoV-2 negative samples

At OUH negative samples were selected from stored pre-pandemic respiratory samples. These had initially been tested with either GeneXpert Flu/RSV (Cepheid) or the BioFire FilmArray Respiratory panel 2.0 (BioMérieux), and were purposefully chosen to include samples with a range of other respiratory pathogens. Over 90% of samples were collected between October-December 2019, but those containing non-SARS-CoV-2 seasonal coronaviruses were used up to a collection data of 10th March 2020 to increase the number available. At STH negatives samples were selected from among those submitted for SARS-CoV-2 testing.

#### RNA extraction

For samples originating from OUH, RNA extraction used the QIAsymphony SP instrument with the DSP Virus/Pathogen Kit and the Complex200_OBL_V4_DSP protocol (Qiagen^12^). 200µl transport medium was added to 430 μl Off Board Lysis buffer then incubated at 68°C for 15 minutes. The whole volume was used for extraction and RNA was eluted in 60µl, then immediately transferred to aliquots and stored at -80°C.

For samples originating from STH, RNA extraction used the MagNA Pure96 instrument with the MagNA Pure 96 DNA and Viral NA Small Volume Kit (Roche). 200µl of transport medium was taken into the extraction and RNA eluted in 100µl before storage at -80°C.

#### Comparator RT-PCR

Comparison RT-PCR assays were undertaken contemporaneously with LamPORE. For samples originating from OUH, the comparator RT-PCR was the RealStar SARS-CoV-2 RT-PCR assay (Altona Diagnostics) on a Qiagen Rotor-Gene Q RT-PCR cycler. This assay takes 10µl RNA input into a single multiplex reaction targeting the SARS-CoV-2 envelope and spike genes, plus an internal control. Samples were analysed in runs including a no template control (NTC), internal quality control (supplied with the RT-PCR kit) and positive control (synthetic genomic SARS-CoV-2 RNA, Twist Bioscience). Results were reported as per OUH criteria for clinical specimens:

##### Positive

Either SARS-CoV-2 target detected at a Ct value ≤37

##### Negative

Neither SARS-CoV-2 targets detected at any Ct value, and sample internal control +/- 3 of the NTC

##### Invalid

Any other result (retested in routine practice, but invalid samples not used for the evaluation)

For samples originating from STH, an in-house RT-PCR assay based on Corman et al.^10^,^11^ provided the comparator, run on an Applied Biosystem 7500 Real Time PCR system. The assay takes 6µl RNA input into a multiplex reaction targeting SARS-CoV-2 E and RdRp genes, plus human RNAse P as an internal control^13^. Results were reported as per STH criteria for clinical specimens:

##### Positive

Either SARS-CoV-2 target detected at a Ct value ≤37.5. Any Ct value single target>37.5 is handled using the referenced algorithm^11^

##### Negative

Neither SARS-CoV-2 target detected and no evidence of inhibition of the internal control

##### Invalid

Internal control not detectable (invalid samples not used for LamPORE evaluation)

LamPORE was performed on one aliquot of RNA, with simultaneous SARS-CoV-2 RT-PCR on another aliquot of the same extract, which was used as the reference. Only samples with a valid SARS-CoV-2 RT-PCR result in keeping with the expected positive or negative categorisation were included in the comparison (10 pre-pandemic negative samples were not used as they had invalid results, and 2 previously positive samples were not used as they had invalid/negative results on repeat testing).

#### Replicates

To assess the reproducibility of the assay, LamPORE replicates were performed on aliquots of the same RNA extract, thawed just prior to analysis. Results from the first replicate were used to report overall diagnostic sensitivity and specificity. To ensure that RT-PCR and LamPORE results were comparable between OUH and STH, a subset of SARS-CoV-2 positive and negative samples were exchanged between sites, with both LamPORE and comparator RT-PCR repeated.

#### Statistical analysis

Results were analysed using R version 3.5.0, with exact binomial confidence intervals calculated for proportions. The initial LamPORE replicates were used to derive estimates of sensitivity and specificity, with second replicates used to estimate LamPORE reproducibility. The full dataset is available in the supplementary material.

## Results

### Limit of Detection

Using spiked samples spiked with cultured virus, LamPORE had a limit of detection of 7-10 SARS-CoV-2 genome copies/µl of extracted RNA or 140-200 copies per 20µl reaction, detecting 24/25 (96%) samples in this range (**table 1**). With the RNA extraction protocol used, this would correspond to a concentration of 2,500-3,600 SARS-CoV-2 genome copies/ml of sample. Although LamPORE did not consistently detect spiked samples at concentrations below this, it was positive in 15/21 (71%) samples at the highest dilution tested, 3-4 genome copies/µl of extracted RNA or 50-70 copies per 20µl reaction, equivalent to 900-1,250 copies/ml of sample.

**Table 1.**
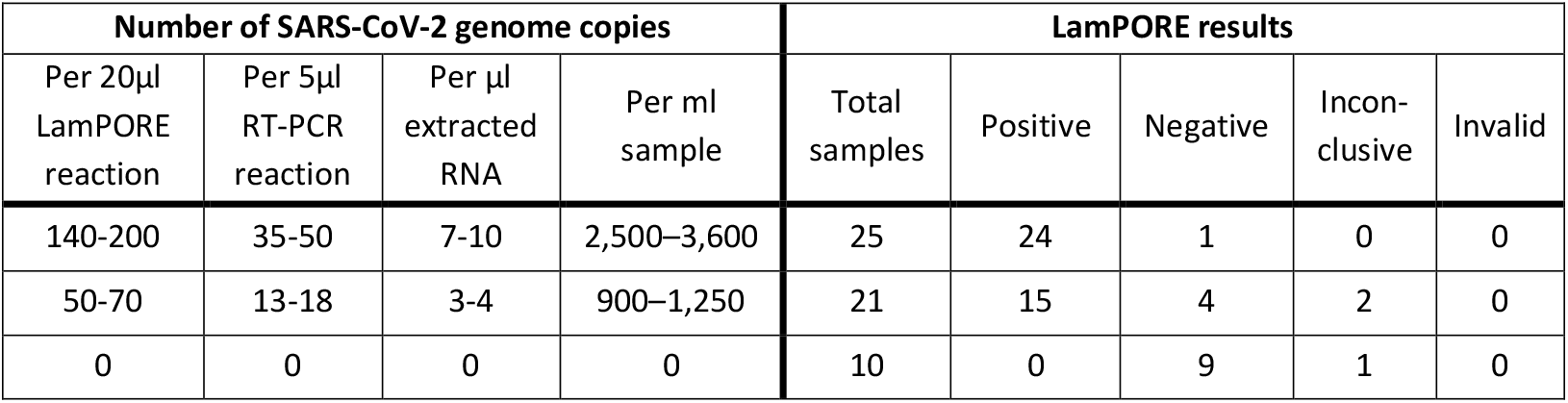
Limit of detection of LamPORE using spiked samples. Copies/RT-PCR reaction is calculated for the comparator CDC NS1 RT-PCR assay using 5µl RNA input volume. Copies/ml sample applies to the extraction method used here, in which RNA from 140µl sample was eluted in 50µl.

| Number of SARS-CoV-2 genome copies |  |  |  | LamPore results |  |  |  |  |
| --- | --- | --- | --- | --- | --- | --- | --- | --- |
| Per 20μl<br>LamPore<br>reaction | Per 5μl<br>RT-PCR<br>reaction | Per μl<br>extracted<br>RNA | Per ml<br>sample | Total<br>samples | Positive | Negative | Incon-<br>clusive | Invalid |
| 140-200 | 35-50 | 7-10 | 2,500–3,600 | 25 | 24 | 1 | 0 | 0 |
| 50-70 | 13-18 | 3-4 | 900–1,250 | 21 | 15 | 4 | 2 | 0 |
| 0 | 0 | 0 | 0 | 10 | 0 | 9 | 1 | 0 |

### Performance of LamPORE

Diagnostic performance of LamPORE was assessed using 514 stored nose and throat swabs, 400 from OUH and 114 from STH (details in **table S1**). Sixty cross-site replicates demonstrated good correlation between RT-PCR Ct values for E gene targets at both sites despite different assays being used at OUH and STH, so this was used as the reference Ct (**figure S1**).

Among 229 RT-PCR-positive samples tested by LamPORE, 226 were reported positive and 2 were reported negative, giving and overall diagnostic sensitivity of 99.1% (226/228 [95%CI 96.9-99.9%])(**table 2**). All valid samples at Ct values of 34.9 or lower were positive by LamPORE (**table 3**). Considering performance at lower viral loads, 7/9 samples with Ct ≥35 were positive and 22/22 of those with Ct values between 30 and 34.9 were positive. Both false negative samples by LamPORE had Ct values ≥38, and one of these was positive by LamPORE on repeat testing (**table S2**). The one RT-PCR positive sample that was invalid on initial LamPORE testing was correctly positive when repeated.

**Table 2.** Clinical diagnostic performance of LamPORE versus RT-PCR.

| RT-PCR<br>result | LamPORE result |  |  |  |  |
| --- | --- | --- | --- | --- | --- |
|  | Positive | Negative | Inconclusive | Invalid | Total |
| Positive | 226 | 2 | 0 | 1 | 229 |
| Negative | 1 | 278 | 3 | 3 | 285 |
| Total | 227 | 280 | 3 | 4 | 514 |

**Table 3.**
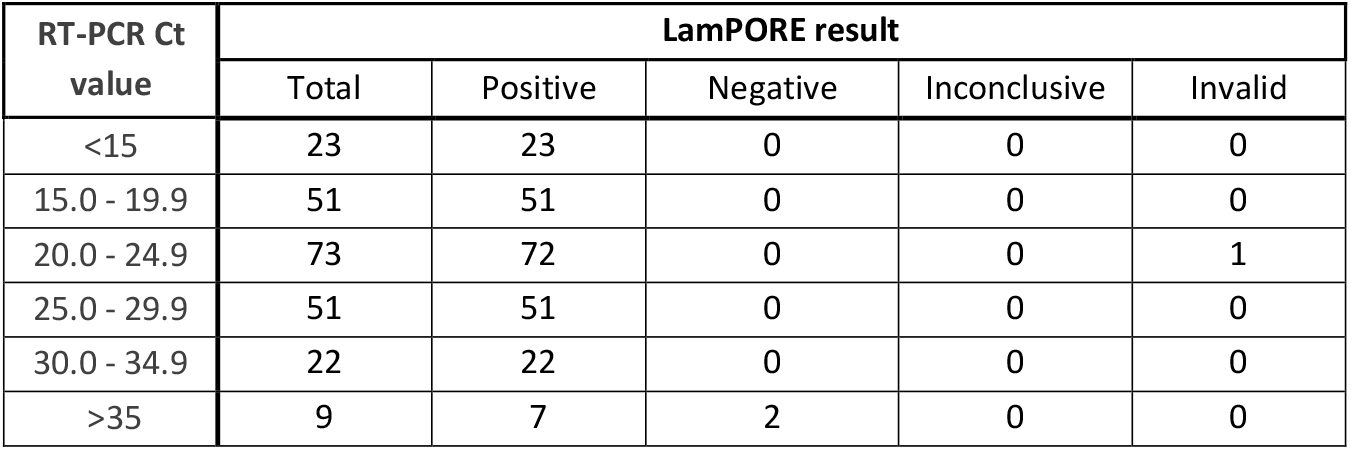
Performance of LamPORE on SARS-CoV-2 positive samples by RT-PCR E gene Ct value.

| RT-PCR Ct value | LamPORE result |  |  |  |  |
| --- | --- | --- | --- | --- | --- |
|  | Total | Positive | Negative | Inconclusive | Invalid |
| <15 | 23 | 23 | 0 | 0 | 0 |
| 15.0 - 19.9 | 51 | 51 | 0 | 0 | 0 |
| 20.0 - 24.9 | 73 | 72 | 0 | 0 | 1 |
| 25.0 - 29.9 | 51 | 51 | 0 | 0 | 0 |
| 30.0 - 34.9 | 22 | 22 | 0 | 0 | 0 |
| >35 | 9 | 7 | 2 | 0 | 0 |

Of 285 RT-PCR-negative samples, 278 were negative and one was positive by LamPORE, giving an overall diagnostic specificity of 99.6% (278/279 [98.0-100.0%]) (**table 2**). The false positive was a pre-pandemic respiratory sample that was also positive for adenovirus, and which had 2,419 SARS-CoV-2 reads detected. However, this sample was negative on repeat LamPORE testing (**figure S2**). Six RT-PCR negative samples gave indeterminate results (three invalid, three inconclusive), of which four were correctly negative on repeat testing, one remained invalid, and one was not retested. Overall, among both RT-PCR-positive and negative samples, 1.4% (7/514 [0.5–2.9]) produced an indeterminate result on first testing.

Another respiratory pathogen was detected by multiplex RT-PCR in 153 negative samples, including 43 with rhinovirus, 38 with RSV, 33 with influenza, and 24 with seasonal coronaviruses (nine HKU1, seven NL63, seven OC43, and one 229E). Overall, there was no evidence that the presence of any other respiratory pathogen was associated with false positive results, or greater numbers of reads assigned to SARS-CoV-2 targets (**figure S2**).

As well as the categorical result produced by the LamPORE reporting algorithm, RT-PCR results were compared with the number of reads assigned by LamPORE to SARS-CoV-2 targets (**figure 1**). This showed that the pre-specified cut-off of ≥50 for a positive result was optimal, with any cut-off in the range of 25-182 producing a maximal Youden index (sensitivity+specificity-1) of 0.988. As the rate at which reads are detected becomes roughly constant within a few minutes of sequencing, the effect of a sequencing run longer or shorter than 60 minutes can be inferred. All samples reported positive by LamPORE had >180 SARS-CoV-2 reads detected, so would have been positive after 30 minutes of sequencing, at which point there would also have been no increase in indeterminate results. Conversely, extending the sequencing duration with the same diagnostic thresholds would not have allowed detection of either of the two false negative samples without producing large numbers of false positives.

**Figure 1.**
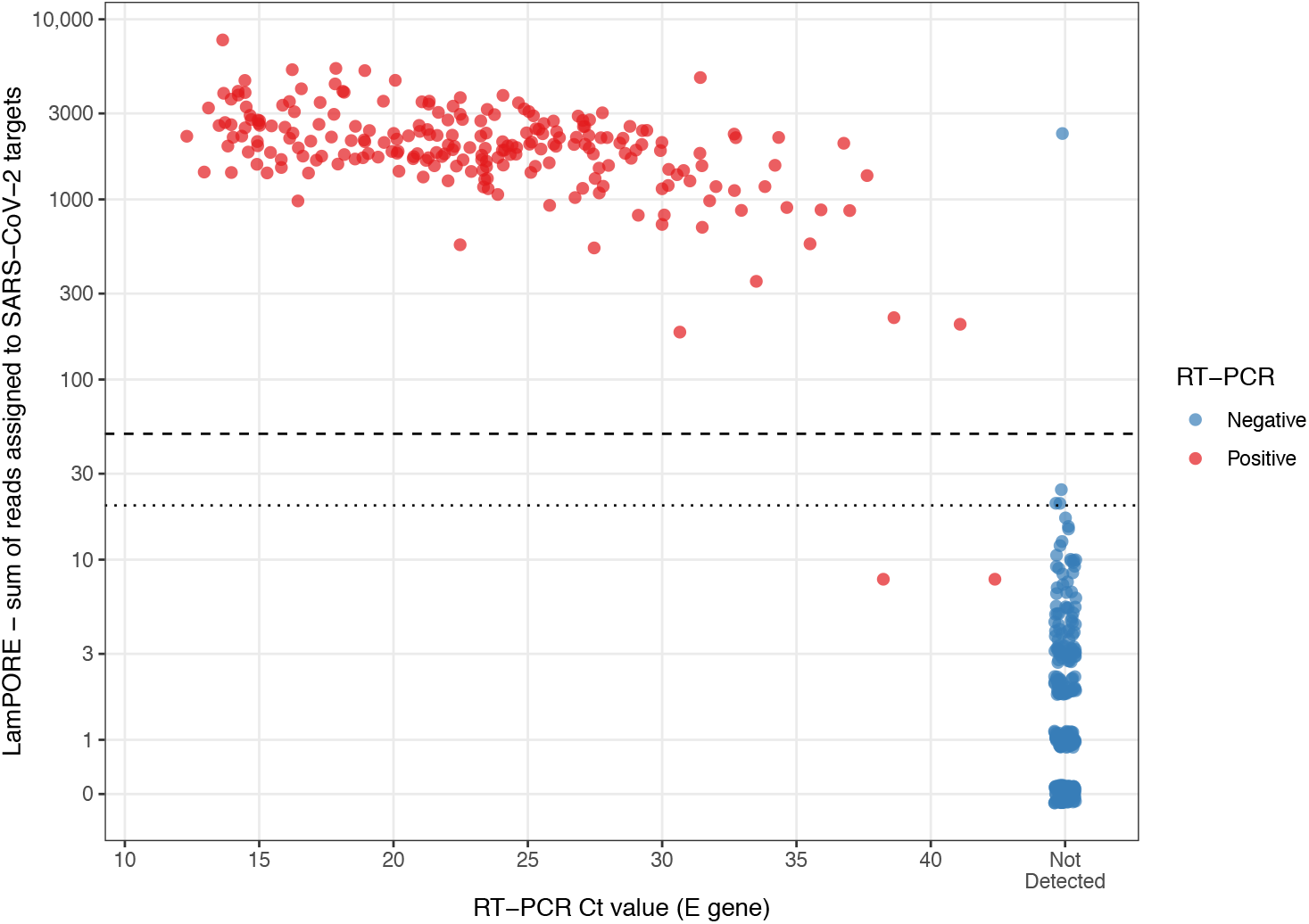
Total number of reads assigned to SARS-CoV-2 targets by LamPORE versus RT-PCR E gene Ct value. Dashed line is threshold for positive result (≥50 reads), dotted line is threshold for inconclusive result (≥20 reads). Invalid samples not plotted.

### Reproducibility

LamPORE was repeated on 494 samples and produced identical results in 478, giving an overall reproducibility of 96.8% (478/494 [94.8-98.1%])(**tables 4 and S3**). In four samples (0.8%) with discrepant LamPORE results, the same sample switched between Negative and Positive. In the other 12 discrepant samples LamPORE replicates included one indeterminate result. All 90 cross-site LamPORE replicates performed between Oxford and Sheffield were concordant (60 RT-PCR/LamPORE positive and 30 RT-PCR/LamPORE-negative).

**Table 4.**
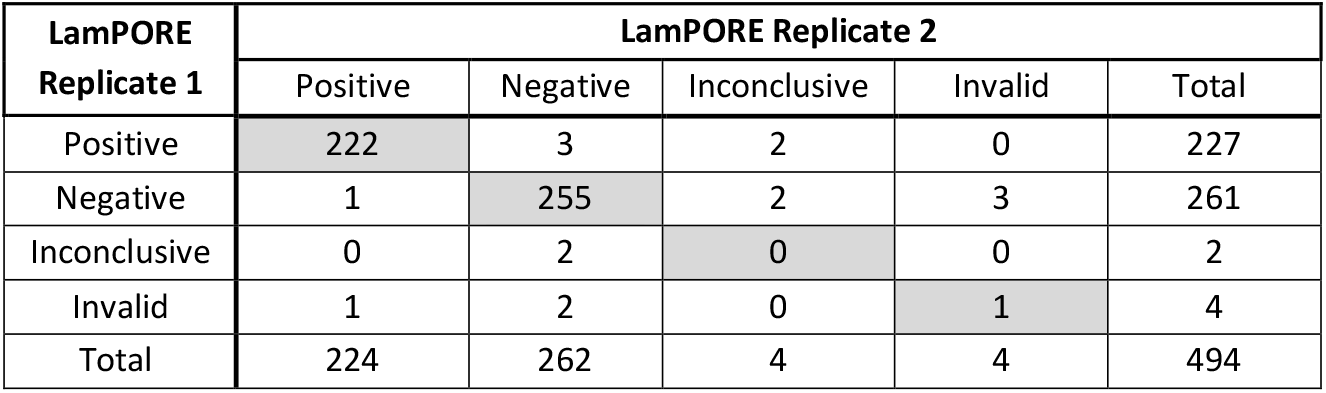
Reproducibility of LamPORE on aliquots of the same RNA extract.

| LamPORE Replicate 1 | LamPORE Replicate 2 |  |  |  |  |
| --- | --- | --- | --- | --- | --- |
|  | Positive | Negative | Inconclusive | Invalid | Total |
| Positive | 222 | 3 | 2 | 0 | 227 |
| Negative | 1 | 255 | 2 | 3 | 261 |
| Inconclusive | 0 | 2 | 0 | 0 | 2 |
| Invalid | 1 | 2 | 0 | 1 | 4 |
| Total | 224 | 262 | 4 | 4 | 494 |

## Discussion

In this evaluation we found that LamPORE had a high diagnostic sensitivity (99.1%) compared to reference RT-PCR in our clinical sample set, consistent with initial development work^5^. The limit of detection of LamPORE, at 7-10 genome copies/µl of extracted RNA, was somewhat higher than the 2 copies/µl achievable with high-performance RT-PCR^14^, but this did not correspond to a significant loss of diagnostic sensitivity in the clinical samples. Our spiking experiments used an extraction in which RNA from 140µl transport medium was eluted in 50µl, which is typical of commonly used protocols. However, commercially available extraction methods can use input volumes of 1ml or more of transport medium, potentially meaning that LamPORE with an input volume of 1ml could have the same limit of detection as RT-PCR with an input volume of 140-200µl. This assumes minimal increase in assay inhibition at higher input volumes, which is currently being evaluated. LamPORE also showed high diagnostic specificity (99.6%) and reproducibility (96.8%), both within and across sites, supporting its practical use for high-throughput testing in a low-prevalence population.

Although no clinical metadata are available for the samples used in this evaluation, they will mainly have been derived from patients with acute symptomatic infection, often requiring admission to hospital, as testing was mainly limited to this group during the first wave of infection. The distribution of Ct values may be higher in a population with more mild or asymptomatic infection, and would be markedly higher among those who remain RT-PCR positive weeks after recovering from acute infection^15,16^. Our data suggest that LamPORE is most likely to miss weak positive samples with Ct values above 35, so could have had lower diagnostic sensitivity if tested in such groups. However, this may not be a significant practical disadvantage, as although weak positives have some value for contact tracing they are likely to come from individuals with low infectious potential^17^.

Our evaluation has several limitations. It was conducted after the first wave of COVID-19 in the UK, when there were few incident cases, so we were unable to prospectively collect samples and instead relied on frozen transport media, which could differ from fresh material. Positives were defined by a positive RT-PCR at the time of initial sample collection and by repeat positive RT-PCR simultaneously with LamPORE, but although RT-PCR is used as a reference test for SARS-CoV-2, there are many reports of its suboptimal sensitivity in clinical infection^18^.

This early evaluation of LamPORE compared its performance against RT-PCR using extracted RNA, as this is the standard material used for detection of SARS-CoV-2. However, the requirement for viral inactivation and RNA extraction in a clinical laboratory produces bottlenecks that mitigate the potential benefit of LamPORE for high-throughput or mobile testing. LAMP reactions are reported to be more robust than RT-PCR to inhibitors present in clinical samples so may have superior performance with extraction-free protocols^19,20^. This could greatly streamline the workflow, but further evaluation is required. We also did not evaluate how the throughput and turnaround time of LamPORE would compare to RT-PCR during routine use in a clinical laboratory or centralised testing centre. Using LamPORE for high-throughput testing of tens or hundreds of thousands of samples per day would be dependent on an streamlined workflow, including automated sample handling and integration with laboratory information management systems.

In conclusion, we show that LamPORE on extracted RNA offers a promising method of high-throughput SARS-CoV-2 testing, and could be of much broader use if shown to work with extraction-free methods of sample preparation and automated workflows.

## Data Availability

Primary data will be available in the supplementary material

## Acknowledgements

We are grateful to all the clinical microbiology/virology staff at OUH and STH who helped to process the specimens used in this evaluation, and to Dr Kevin Bewley, PHE Porton Down, for providing the cultured virus.

## Ethics

The process for collection of the donated saliva was approved by the PHE Research Ethics and Governance Group. The protocol for the use of stored clinical samples at OUH and STH was reviewed by the Joint Research Office of OUH and the University of Oxford (our Institutional Review Board), and it was determined that the activity constituted service evaluation and service development. As such, it required neither sponsorship nor research ethics review.

## Supplementary Figures

**Figure S1.**
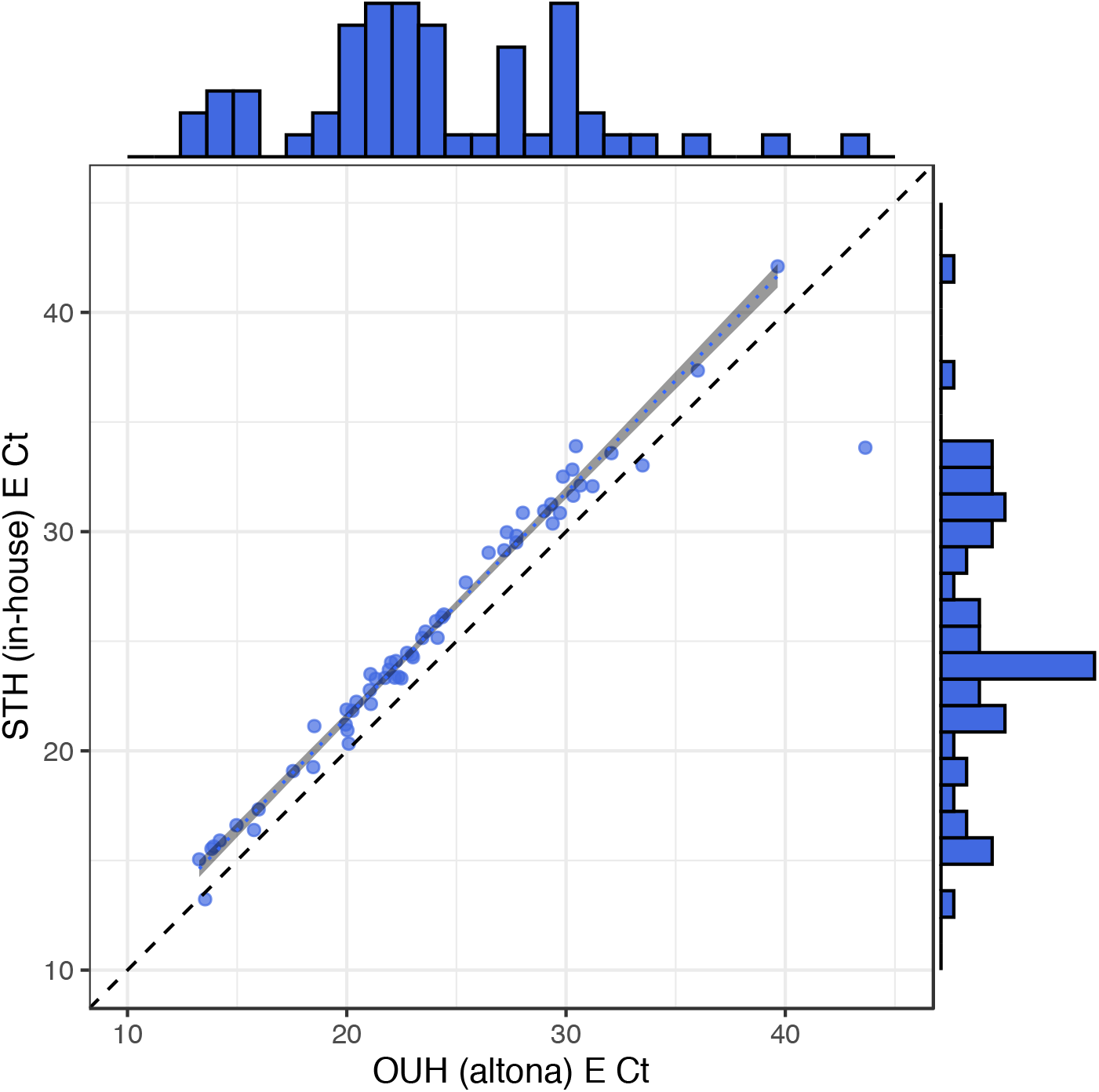
Comparison of E gene Ct values for 60 SARS-CoV-2 positive cross-site replicates between OUH and STH. Ct values at OUH were approximately one cycle lower than at STH, in part because of greater RNA input volume (10µl at OUH vs 6µl at STH). This line of best fit is plotted with shaded area representing 95% confidence intervals and excludes the single outlier.Organism Detected by PCR

**Figure S2.**
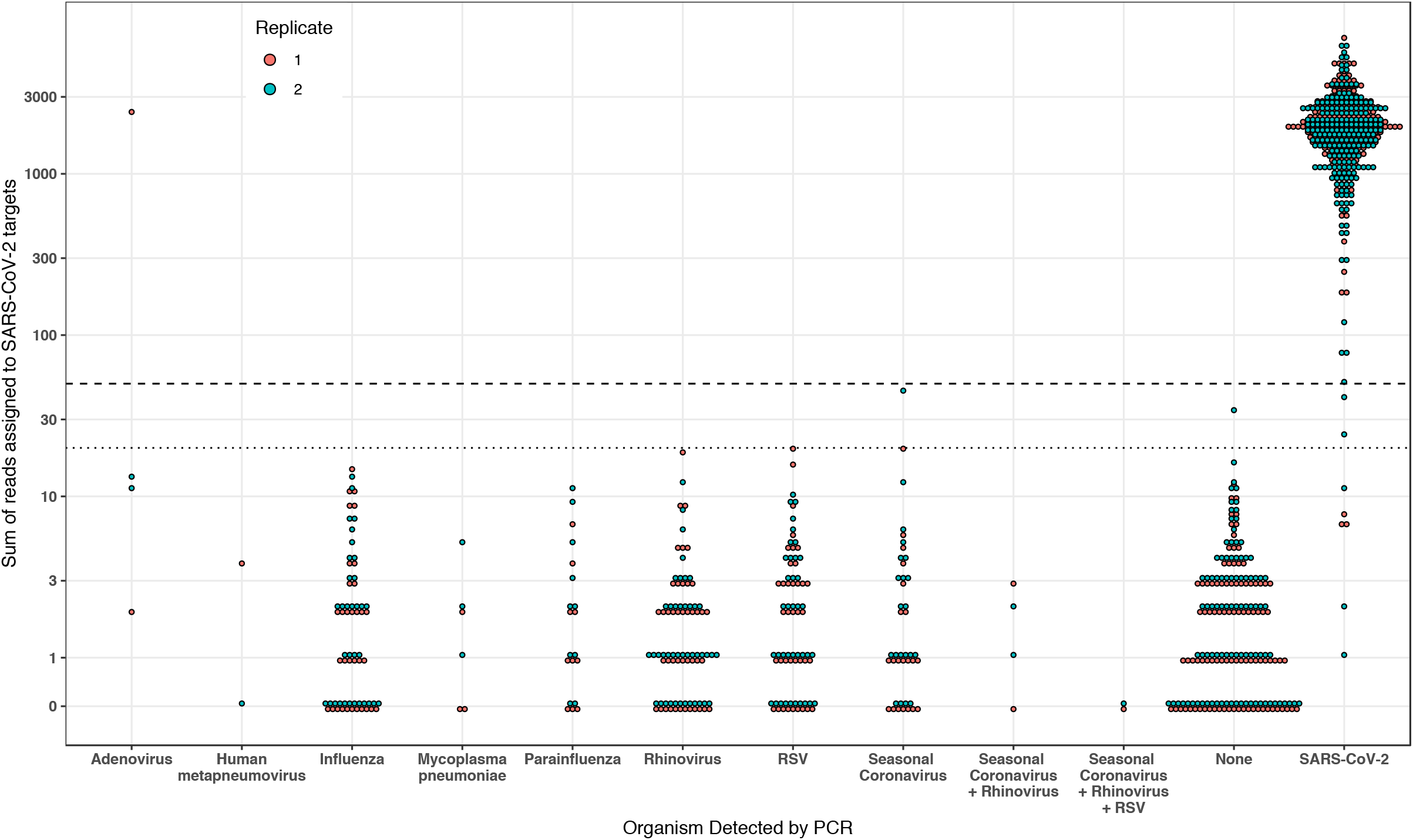
Analytical specificity of LamPORE in samples positive for a range of respiratory pathogens. Data for both LamPORE replicates is shown.Dashed line is threshold for positive result (≥50 reads) and dotted line is threshold for inconclusive result (≥20 reads). Invalid samples are plotted.

## Supplementary Tables

**Table S1.** Sources of samples used for retrospective clinical evaluation.

| Sample type<br>(origin site – site replicated) | Number of RT-PCR<br>positive samples | Number of RT-PCR<br>negative samples |
| --- | --- | --- |
| OUH – OUH | 120 | 235 |
| OUH – STH | 30 | 15 |
| STH – OUH | 30 | 15 |
| STH – STH | 49 | 0 |
| STH (not replicated) | 0 | 20 |

**Table S2.** LamPORE results for false negatives in replicates 1 and 2.

| Sample | LamPORE replicate 1 | LamPORE replicate 2 | E gene Ct |
| --- | --- | --- | --- |
| False negative 1 | Negative | Negative | 42.1 |
| False negative 2 | Negative | Positive | 38.1 |
| False negative 3 | Positive | Negative | 37.0 |
| False negative 4 | Positive | Negative | 22.9 |

**Table S3.**
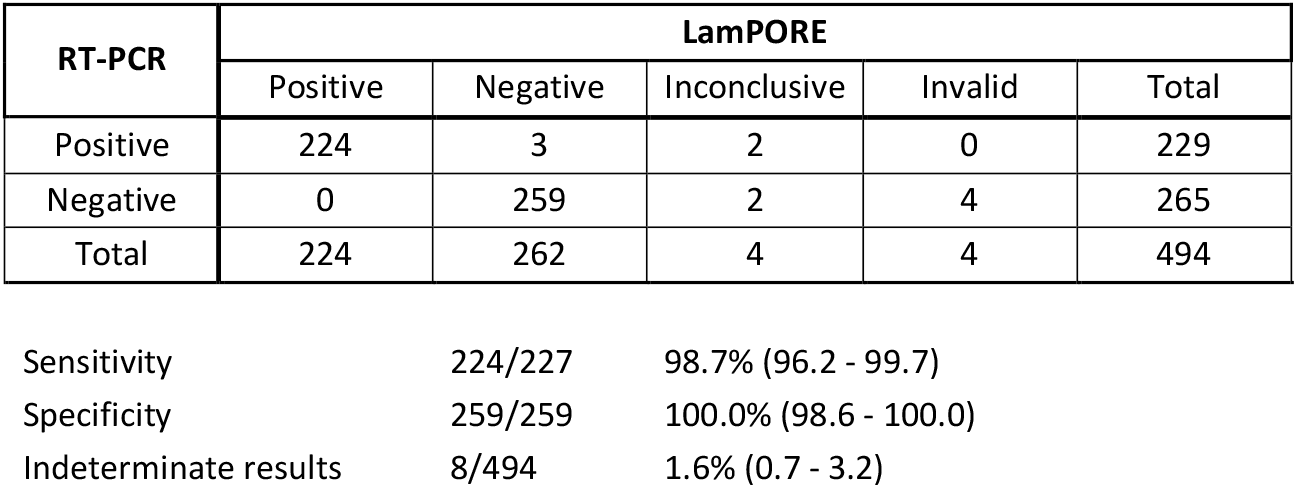
Performance versus RT-PCR for 2^nd^ LamPORE replicates (The performance of the 1^st^replicate was used as the primary outcome).

| RT-PCR | LamPORE |  |  |  |  |
| --- | --- | --- | --- | --- | --- |
|  | Positive | Negative | Inconclusive | Invalid | Total |
| Positive | 224 | 3 | 2 | 0 | 229 |
| Negative | 0 | 259 | 2 | 4 | 265 |
| Total | 224 | 262 | 4 | 4 | 494 |

|  |  |  |
| --- | --- | --- |
| Sensitivity | 224/227 | 98.7% (96.2 - 99.7) |
| Specificity | 259/259 | 100.0% (98.6 - 100.0) |
| Indeterminate results | 8/494 | 1.6% (0.7 - 3.2) |

## References

1. Grassly, N. C. et al. Comparison of molecular testing strategies for COVID-19 control: a mathematical modelling study. The Lancet Infectious Diseases (2020). doi:10.1016/S1473-3099(20)30630-7

2. Peto, J. et al. Weekly COVID-19 testing with household quarantine and contact tracing is feasible and would probably end the epidemic. R Soc Open Sci 7, 200915 (2020).

3. Fowler, V. L. et al. A reverse-transcription loop-mediated isothermal amplification (RT-LAMP) assay for the rapid detection of SARS-CoV-2 within nasopharyngeal and oropharyngeal swabs at Hampshire Hospitals NHS Foundation Trust. medRxiv 2020.06.30.20142935 (2020). doi:10.1101/2020.06.30.20142935

4. Rodriguez-Manzano, J. et al. A handheld point-of-care system for rapid detection of SARS-CoV-2 in under 20 minutes. medRxiv 2020.06.29.20142349 (2020). doi:10.1101/2020.06.29.20142349

5. James, P. et al. LamPORE: rapid, accurate and highly scalable molecular screening for SARS-CoV-2 infection, based on nanopore sequencing. medRxiv 2020.08.07.20161737 (2020). doi:10.1101/2020.08.07.20161737

6. Schmid-Burgk, J. L. et al. LAMP-Seq: Population-Scale COVID-19 Diagnostics Using Combinatorial Barcoding. bioRxiv 25, 2431 (2020).

7. Thi, V. L. D. et al. A colorimetric RT-LAMP assay and LAMP-sequencing for detecting SARS-CoV-2 RNA in clinical samples. Sci Transl Med 12, eabc7075 (2020).

8. CDC 2019 Novel Coronavirus (nCoV) Real-Time RT-PCR Diagnostic Panel - Instructions for Use. fda.gov Available at: https://www.fda.gov/media/134922/download. (Accessed: 10 September 2020)

9. guidance-and-sop-covid-19-virus-testing-in-nhs-laboratories-v1.pdf. england.nhs.uk Available at: https://www.england.nhs.uk/coronavirus/wp-content/uploads/sites/52/2020/03/guidance-and-sop-covid-19-virus-testing-in-nhs-laboratories-v1.pdf. (Accessed: 10 September 2020)

10. Corman, V. M. et al. Detection of 2019 novel coronavirus (2019-nCoV) by real-time RT-PCR. Eurosurveillance 25, 2431 (2020).

11. Colton, H. et al. Improved sensitivity using a dual target, E and RdRp assay for the diagnosis of SARS-CoV-2 infection: Experience at a large NHS Foundation Trust in the UK. J. Infect. (2020). doi:10.1016/j.jinf.2020.05.061

12. QIAsymphony SP Protocol Sheet Complex200_OBL_V4_DSP protocol - QIAGEN. qiagen.com Available at: https://www.qiagen.com/gb/resources/resourcedetail?id=02c3171f-9867-43eb-b48b-135afe95b29e&lang=en. (Accessed: 10 September 2020)

13. Thurman, K. A., Warner, A. K., Cowart, K. C., Benitez, A. J. & Winchell, J.M. Detection of Mycoplasma pneumoniae, Chlamydia pneumoniae, and Legionella spp. in clinical specimens using a single-tube multiplex real-time PCR assay. Diagnostic Microbiology and Infectious Disease 70, 1–9 (2011).

14. SARS-CoV-2 molecular assay evaluation: results - FIND. SARS-CoV-2 molecular assay evaluation: results - FIND doi:10.1002/14651858.CD013

15. Singanayagam, A. et al. Duration of infectiousness and correlation with RT-PCR cycle threshold values in cases of COVID-19, England, January to May 2020. Eurosurveillance 25, 465 (2020).

16. Rao, S. N., Manissero, D., Steele, V. R. & Pareja, J. A Systematic Review of the Clinical Utility of Cycle Threshold Values in the Context of COVID-19. Infect Dis Ther 9, 573– 586 (2020).

17. Jefferson, T., Spencer, E., Brassey, J. & Heneghan, C. Viral cultures for COVID-19 infectivity assessment. Systematic review. medRxiv 2020.08.04.20167932 (2020). doi:10.1101/2020.08.04.20167932

18. Woloshin, S., Patel, N. & Kesselheim, A. S. False Negative Tests for SARS-CoV-2 Infection - Challenges and Implications. N. Engl. J. Med. 383, e38 (2020).

19. Kaneko, H., Kawana, T., Fukushima, E. & Suzutani, T. Tolerance of loop-mediated isothermal amplification to a culture medium and biological substances. Journal of Biochemical and Biophysical Methods 70, 499–501 (2007).

20. Tani, H. et al. Technique for quantitative detection of specific DNA sequences using alternately binding quenching probe competitive assay combined with loop-mediated isothermal amplification. Anal. Chem. 79, 5608–5613 (2007).

